# Moment-to-moment fluctuations of hemodynamic responses in posterior default mode networks differentially predict level of attentional lapses in adolescents with Attention-Deficit/Hyperactivity Disorder

**DOI:** 10.1101/2022.01.31.22270169

**Authors:** Lin Sørensen, Yu Sun Chung, Sabin Khadka, Michael C. Stevens

## Abstract

**Background:** The neurobiological underpinnings of the characteristically higher intra-individual variability of reaction times (IIVRT) in patients with ADHD remain poorly understood. The aim of the current study was to characterize the role of the default mode and other canonical brain networks measured by functional magnetic resonance imaging (fMRI) to task performance fluctuations measured by IIVRT. To our knowledge, no prior fMRI study has shown the involvement of posterior default mode network (DMN) in ADHD IIVRT. We expected that moment-to-moment fluctuations in hemodynamic responses in posterior DMN would predict higher IIVRT in ADHD.

**Methods:** Adolescents (12 to 19 years old) with ADHD (n= 55) and healthy controls (n= 55) performed a fMRI Go/NoGo task. Whole-brain independent component analysis (ICA) segregated hemodynamic responses into functional brain networks, then further decomposed into individual trial-specific estimates of hemodynamic response amplitude. Mean and variability metrics of these amplitudes were tested in stepwise linear regression analyses to identify which functional brain networks predicted high IIVRT.

**Results:** As hypothesized, variability in hemodynamic responses in posterior DMN regions predicted level of IIVRT. In posterior cingulate cortex this variability predicted higher IIVRT only in ADHD, whereas in precuneus variability in hemodynamic responses predicted lower IIVRT. Average hemodynamic responses in a bilateral superior temporal cortex network predicted higher IIVRT only in ADHD.

**Conclusion:** Our findings suggest that estimating variability in hemodynamic responses is crucial to understand the involvement of the intrinsic default mode in attentional lapses in ADHD. The parcellation into subnetworks showed the differentiating role of default mode in attentional lapses in ADHD.

## Introduction

Attention-Deficit/Hyperactivity Disorder (ADHD) is a neurodevelopmental disorder found in about 5% of children and adolescents (1). Consistent with ADHD’s age-inappropriate elevated symptoms of inattention, lapses in attention occur on cognitive tests in ADHD-diagnosed persons more frequently than in non-ADHD controls (2-5) (Hedges’ *g* effect size = 0.76, or “large”) (3). In research studies, attentional lapses typically are operationalized as moment-to-moment fluctuations in task-performance measured with intra-individual reaction time variability (IIVRT), and sometimes as longer mean reaction times (RTs) (2, 4-6). IIVRT can occur in many different cognitive contexts (7) (e.g., in paradigms that measure attention or cognitive control (5)), but appears to be unique from other shared performance characteristics like response speed (3, 8). IIVRT is a candidate endophenotype for ADHD. It not only has a genetic basis (9, 10) and appears to be a lifetime trait (3), IIVRT predicts academic performance in children with ADHD (11), can differentiate ADHD from autism (8, 12), and there is evidence suggesting IIVRT abnormalities are attenuated by psychostimulant ADHD medications (3). Despite these numerous indicators that IIVRT is an important ADHD biomarker, its exact neurobiological underpinnings are still poorly understood.

One dominant hypothesis is that attentional lapses stem from spontaneous activity within the brain’s intrinsic default mode network (DMN) that interrupts goal-directed “task-positive” brain activation (6, 13, 14). The DMN is a collection of network functional hubs (medial prefrontal cortex (mPFC), posterior cingulate cortex (PCC) and precuneus) that seem to shift back-and-forth from a “default” state with high baseline activity to an active cognitively demanding state (13). Performing externally-oriented, attention-demanding tasks normally will lead to suppression of DMN spontaneous activity (6, 13, 14). In ADHD, it has been proposed that spontaneous activity within DMN regions might persist to interrupt attention processing, causing RT fluctuations and high IIVRT (15, 16). In resting-state fMRI studies, DMN activation has been shown to be abnormally higher in ADHD (17 – 21), while the typical inverse correlation between DMN activity and task-based cognitive control network hubs appears weaker and to suffer from a maturational lag in ADHD (17-21). ADHD IIVRT is associated with mean hemodynamic responses (HRs) in the anterior DMN regions of mPFC during working memory processing (22) and during attention oddball task processing (23). Some evidence also links behavioral IIVRT to variability in the strength of the hemodynamic response itself within anterior DMN brain regions (24). Importantly, this latter study was the first to show IIVRT was not simply the result of weaker or lower brain activity, but more specifically related to the inconsistency of response to attention-eliciting stimuli on an oddball attention task.

Although such studies have greatly advanced our understanding of ADHD neural dysfunction underlying IIVRT, the posterior DMN regions have not yet been linked to IIVRT in ADHD. This absence of evidence is meaningful, as in some ways the posterior DMN represents a more likely, logical neurobiological influence over attention fluctuations. Several studies suggest posterior DMN regions are especially important for the transition between the low-frequency resting-state brain activation and the down-regulation of this fluctuation during task-positive activation of attention networks in the brain (13, 25, 26). Interestingly, in fMRI meta-analyses of Go/NoGo tasks, individuals with ADHD typically show hyperactivation – or what could be interpreted as lower deregulation (27) – of the posterior DMN areas of the precuneus/PCC during motor inhibition (NoGo) (27-29). However, none of those studies showed an association between DMN regions and ADHD IIVRT. High IIVRT in ADHD instead has been associated with mean hemodynamic responses in frontoparietal attention networks (30, 31) and temporal areas (32). Also, attentional lapses as estimated with the ex-gaussian distribution of reaction times (tau) on Go trials was not associated with mean amplitude of hemodynamic responses in any functional brain network (33). Because these latter findings do not strictly comport with the DMN interruption hypothesis, they raise the possibility that the underlying neural mechanism of IIVRT-related ADHD dysfunction might be different, more complex, or more extensive than is currently thought (16). For instance, high IIVRT in ADHD could stem from abnormal motor response preparations and selection involving the pre-motor systems (34), or even temporal processing deficits involving other cortex (2, 23, 32). There is a need to more rigorously interrogate all possibly-relevant brain systems to aid ADHD IIVRT neurobiological modeling-building efforts.

Accordingly, this study’s goal was a) to evaluate task-elicited responsiveness and variability concurrently within all the brain’s major neural systems to determine which are linked to high IIVRT and are dysfunctional in ADHD, while retaining a specific focus on DMN as the leading candidate neural system. We hoped to extend prior findings of IIVRT-related neurobiology primarily learned from attention paradigms by examining a moderate-sized adolescent non-ADHD and ADHD sample who performed a Go/NoGo fMRI task (35). We predicted that Go/NoGo task hemodynamic response amplitude variability in both anterior and posterior DMN hub regions would predict higher ADHD IIVRT (24, 36, 37). Secondary expectations included that mean amplitude would predict similarly as previous Go/NoGo fMRI studies higher IIVRT in pre-motor systems (31, 38) and temporal lobe cortex (2, 23).

## Methods and Materials

### Participants

The sample (*N*=110) consisted of adolescents with ADHD (*n*=55) and healthy controls between 12 and 19 years old (M=15.40, SD=1.77). Experienced clinical staff conducted a diagnostic evaluation using the Schedule for Affective Disorders and Schizophrenia for School-Age Children-Present and Lifetime Version (K-SADS-PL) (39). In the ADHD group, 49 were diagnosed with the DSM-V Combined-Type ADHD, 1 with predominantly inattentive ADHD and 5 with an ADHD in remission. Comorbid disorders were as typically reported (40) in ADHD: Oppositional Defiant Disorder (*n*=10), Conduct Disorder (*n*=4), anxiety and depressive disorders (*n*=6), and substance use disorders (*n*=7). In the control group, 1 had a former depressive disorder and 1 substance use disorder. Full Scale intelligence (IQ) was estimated using the Wechsler Abbreviated Scale of Intelligence (WASI) (41). ADHD participants who took medication followed a 24-hour washout procedure prior to the cognitive and MRI assessments. All participants and/or their parents gave informed consent following the procedures approved by Hartford Hospital’s Institutional Review Board and in accordance with the Helsinki declaration.

### fMRI task and procedure: Go/NoGo task

Participants performed two sessions of an event-related fMRI Go/NoGo task consisting of frequent “X” and infrequent “K” stimuli presented for 50 msec each (35, 42). Each lasted 7:21 min and consisted of 246 trials, in which the “K” stimulus was presented in 16% of the trials with intervals in the range of 10-15 seconds. Participants were instructed to make a speeded button press with their right index finger to rapidly presented “X” (Go) stimuli, but to withhold response to pseudo-randomly interspersed “K” (NoGo) stimuli. The participants first performed a practice trial to ensure they understood instructions. Responses were registered using a fiber-optic MRI-compatible response device (Current Designs, Inc.) and recorded for off-line analyses. Hits and errors were registered as responses occurring within 1,000 msec of an “X” or “K” trial, respectively. IIVRT and mean reaction times were estimated by calculating standard deviations and means for each participant across all Go-trials in the two sessions. The Go/NoGo task was implemented using the E-Prime 2.0 (Psychology Software Tools, Inc.). Stimuli were projected to MRI via a screen visible to the participants by a rear-facing mirror attached to the head coil.

### MRI Scanning Parameters

MR imaging was performed with a 3T Siemens Allegra scanner at the Olin Neuropsychiatric Research Center, Institute of Living at Hartford Hospital. Functional image volumes were collected using gradient-echo echo-planar imaging (EPI) pulse sequence: repetition time (TR)=1500 msec, echo time (TE)=28 msec, flip angle=65°, field of view=24×24 cm, acquisition matrix=64×64, A>>P phase encoding, voxel size=3.4×3.4 mm, slice thickness=5 mm, number of slices=29 (acquired sequentially). Gradient echo fieldmaps: TR=580 msec, TE=7 msec, flip angle=90°, matrix=128×128, A>>P phase encoding, 3 mm slice thickness, MPRAGE T1-weighted images of brain structure: TR=2500 msec, TE=2.74 msec, flip angle=8°, matrix=256×208, 1 mm slice thickness.

### Image Processing

Each of the fMRI time series was realigned to the mid-series volume (43), corrected for slice-timing acquisition differences (44). Spatial distortions were removed when due to inhomogeneity according to fieldmap-based unwarping (45). Signal spikes were removed using AFNI 3Despike (46) and volumes were automatically reoriented to sterotactic space using 3-parameter rigid body realignment. Spatial normalization parameters mapping the T1 to MNI atlas space were applied to each fMRI volume. Each image was written at 3 mm^3^ isotropic voxel resolution and thereafter spatially smoothed with a 6 mm FWHM Gaussian kernel. Variance in time series due to had motion was removed and the data linearly detrended using fMRIB’s ICA-based Xnoiseifier (FIX v1.602 beta) (47).

### Independence component analysis (ICA)

A group-level ICA was performed with subsequent extraction of single-trial estimates and regression as described in Eichele et al. (37). This allowed for analyzing the temporal dynamics of successive task-trials of the Go/NoGo. The ICA functionally segregates individual or small groups of brain regions with unique time courses (48, 49). The preprocessed data were decomposed into 100 components using a high model order ICA with the GIFT toolbox (https://trendscenter.org/software/gift/). Spatial ICA maps were generated with the Infomax algorithm (37, 50). Regression analysis was used to back-construct participant-specific timecourses (51). 32 task-related components were selected for analysis, based on whether or not each had hemodynamic timecourses that correlated with a conventional fMRI model of Go/NoGo task onsets for correct rejections, commission errors, or hits across the sample. For these components, single-trial estimates of the hemodynamic responses were calculated for each participant. The single-trial mean of amplitudes is comparable to simple activation from conventional fMRI time series calculated with GLM analysis. Next, intra-individual standard deviations of single-trial hemodynamic responses across trials were estimated to measure variability in these response amplitudes. Larger standard deviations reflect greater intra-individual trial-to-trial hemodynamic response variability (52).

### Testing the prediction of single-trial amplitude ICs on level of IIVRT

All statistical analyses were performed with the IBM SPSS version 26. Between-group analyses were conducted to investigate effects of ADHD on level of IIVRT, and bivariate correlational analyses to study the relationship between the ICs. Prior to modeling, data outliers were defined by calculating the median absolute deviation (MAD), using a threshold of +/- 3 for calculating the deviation distance from the center in terms (53). Across the 32 ICs of hemodynamic response variability and mean measurements over all types of trials (192 variables), 1.7% outlier cases in average were detected. These infrequent outliers were replaced with *M* +/- 3\**MAD* scores.

To test which functional brain networks predicted level of IIVRT, stepwise (forward) linear regression analyses were conducted. In all models, age, sex, and IQ were included as covariates. All models used IIVRT as the outcome variable and the single-trial mean and variability for each of the 32 ICs and the diagnoses of ADHD as predictors, both as main effects and as interaction effect terms (ICs×ADHD). Three of these models used Correct Reject, False Alarm and Hit hemodynamic response variability for the ICs, while another three used hemodynamic response mean. The stepwise procedure involved conducting multiple linear regression analyses several times, each step removing the variables with weakest correlation with the outcome variable of IIVRT. To maximize predicted variance of each of the models, we focused on the last regression model to reach significance. This last model was bootstrapped as a resampling procedure.

The overall regression models were:

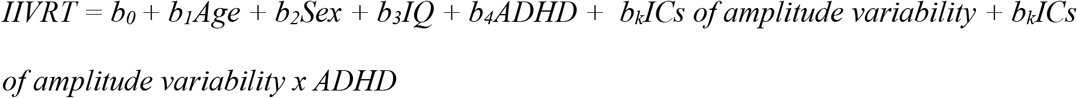

The significance threshold for multiple hypotheses testing was adjusted according to primarily testing the effect of hemodynamic response variability to the three Go/NoGo parameters of Correct Rejections, False alarms, and Hits, respectively, on level of IIVRT in ADHD (*p*<0.017; Bonferroni 0.05/3). For the bootstrapping procedure, *p* <.05 was used. In all the stepwise regression models, the variance inflation factor (VIF) was < 2.

## Results

### fMRI data independent component (IC) maps

Correlation of IC spatial structure with stereotactic atlases found 8 of the 32 ICs examined represented DMN regions (see Figure 1 and Supplemental Table 1). IC 35, IC 39, IC44, IC 53, IC 71, and IC 74 were in posterior DMN. IC 35 and IC 44 showed an intensity peak in the PCC, whereas IC 71 and IC 74 represent dorsal and ventral precuneus, respectively. IC 39 and 53 correlated highly with posterior DMN regions as well. However, IC 53 also included superior parietal lobule on the lateral surface contiguous with precuneus. The PCC timecourses depicted in IC 39 also correlated with mid-cingulate regions, which are not recognized as DMN regions. IC 50 and IC 70 represented anterior DMNs with intensity peaks in the vmPFC, although IC 70 also showed involvement of the PCC. The other ICs represented, among others, a bilateral superior temporal network (IC 15), a right post-motor system network (IC 18) and attention-related networks (see Supplemental Table 1).

**Table 1.** Descriptive information about the groups of adolescents with ADHD and healthy controls.

| Variables | ADHD |  | HC |  | Analysis |  |  |
| --- | --- | --- | --- | --- | --- | --- | --- |
|  | Mean | SD | Mean | SD | *t* | *f*# | *f*## |
| Age | 15.297 | 1.80 | 15.476 | 1.75 | -0.53 |  |  |
| Full-Scale IQ | 105.345 | 13.4649 | 110.628 | 10.0780 | -2.329* |  |  |
| Mean RT | 0.3776 | 0.04069 | 0.3716 | 0.03929 | 0.79 | 0.31 |  |
| IIVRT | 0.1416 | 0.04917 | 0.1058 | 0.03298 | 4.480** | 17.80** | 17.79** |
|  | *n* males | *n* females | *n* males | *n* females |  |  |  |
| Sex | 43 | 12 | 43 | 12 |  |  |  |

Note. \*\* $p < .001$ ; \* $p < .05$ . RT = reaction times; IIVRT = intra-individual variability in reaction times; # = between group effects adjusted for distribution of age, sex, and Full-scale IQ; (df = 1,105); ## = between-group effect adjusted for distribution of Mean RT, age, sex, and Full-scale IQ (df = 1,104).

**Figure 1.**
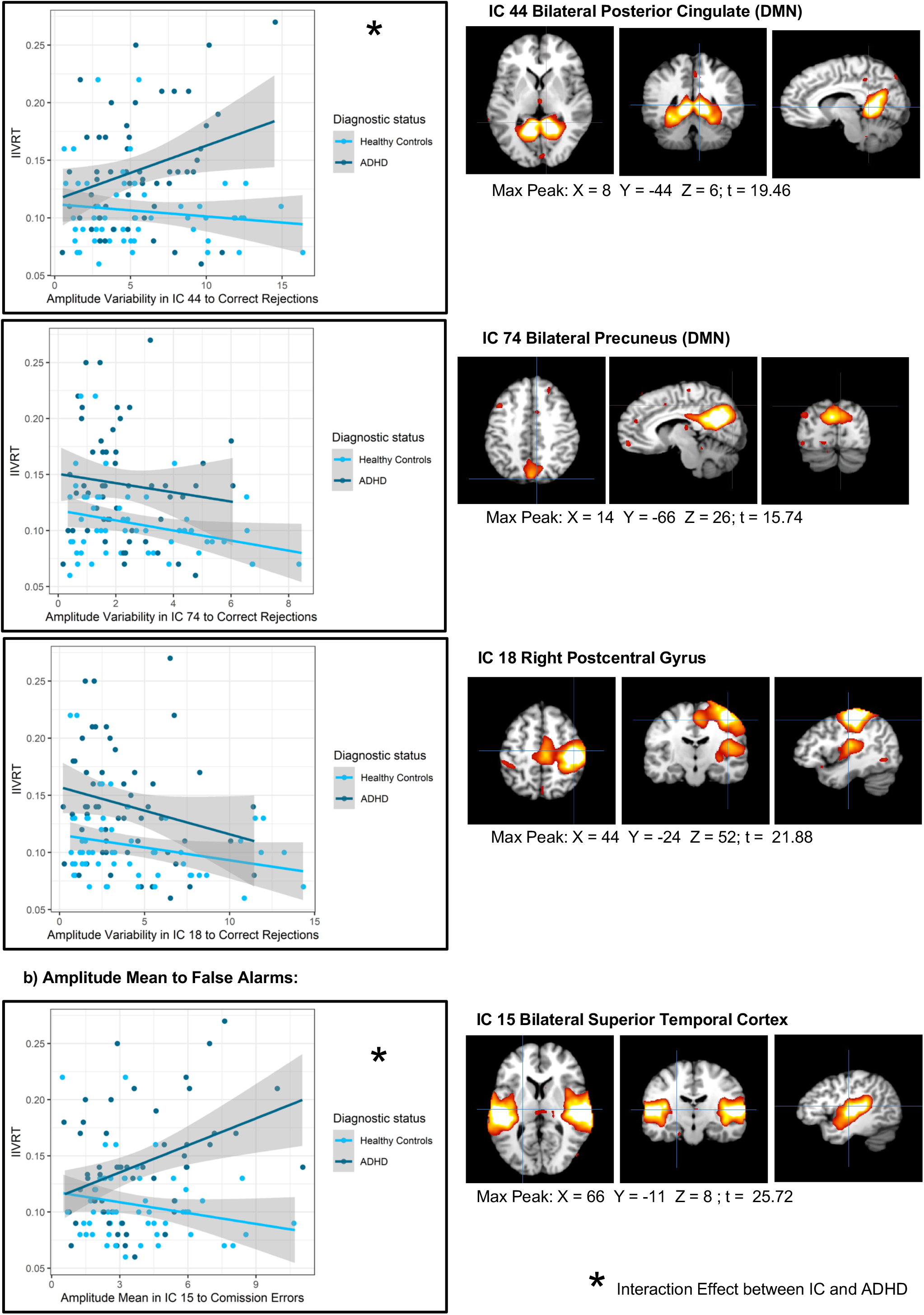
Scatterplots of brain networks involved in the relationship between ADHD, amplitude variability in hemodynamic responses and IIVRT. Note. The X axes includes different ranges of amplitude values due to differences in amplitude level between the different ICs.

### Preliminary descriptive analyses

The group of adolescents with ADHD and healthy controls matched in age and sex distributions, whereas the adolescents with ADHD had lower IQ than the healthy controls (see Table 1). ADHD-diagnosed adolescents had higher IIVRT compared with the healthy controls, after conducting ANCOVAs controlling for effects of age, sex, full-scale IQ, and mean reaction times. Mean reaction time did not differ between study groups. In the ANCOVAs, age (*F*(1,104)=8.25, *p*=0.005) and mean reaction times (*F*(1,104)=10.01, *p*=0.002) covaried with level of IIVRT, whereas sex and full-scale IQ did not. Bivariate correlations showed lower age correlated with higher IIVRT, females showed a higher IIVRT than males, and higher IIVRT correlated with higher mean reaction times (see Supplementary Table 2).

### Variability in hemodynamic responses on level of IIVRT

#### Correct Rejections

The bootstrapped regression model that predicted maximized variance of IIVRT included a significant interaction effect between ADHD and higher variability in hemodynamic responses to Correct Rejections in a bilateral, posterior DMN (IC 44). Higher amplitude variability in IC 44 specifically in ADHD predicted higher IIVRT (see Figure 1 and Table 2). In addition, there were significant main effects of higher variability in hemodynamic responses to correct rejections in a bilateral posterior DMN (IC 74) and a right post-motor system to predict lower IIVRT (IC 18 (see Table 2). ADHD diagnosis and lower age predicted higher IIVRT, whereas sex or IQ did not. Bivariate *post hoc* correlation analyses showed hemodynamic response variability to Correct Rejections in IC 44 correlated positively with hemodynamic response variability to Correct Rejections in IC 74; however, not with hemodynamic response variability to Correct Rejections in IC 18 or with level of IIVRT, mean reaction times, age, sex, or full-scale IQ (see Supplementary Table 3). Variability in hemodynamic responses to Correct Rejections in IC 74 correlated negatively with level of IIVRT and not with hemodynamic response variability to Correct Rejections in IC 18, mean reaction times, age, sex, or full-scale IQ.

**Table 2.** Maximized predicted variance from stepwise linear regression analyses of hemodynamic responses on ADHD IIVRT - the last model with highest explained variance with p<.017 selected.

| Hemodynamic response variability to go/no-go correct rejections (CR): |  |  |  |  |  |  |  |
| --- | --- | --- | --- | --- | --- | --- | --- |
| Adj. $R^2$ = 0.38 | $df$ = 101 | | $F$ = 9.20 | | p <0.001 | | |
|  |  |  |  | Bootstrapping: |  |  |  |
| Predictors | B | $\beta$ | $p$ | $p$ | SE | 95% CI for B | |
| Age | -0.007 | -0.29 | <0.001* | 0.002 | 0.002 | -0.010 | -0.003 |
| Sex | -0.004 | -0.04 | 0.631 | 0.657 | 0.009 | -0.024 | 0.012 |
| Full-scale IQ | 0.000 | -0.08 | 0.287 | 0.324 | 0.000 | -0.001 | 0.000 |
| ADHD | 0.030 | 0.33 | <0.001* | 0.001 | 0.007 | 0.015 | 0.044 |
| CRIC18SD | -0.003 | -0.22 | 0.006* | 0.005 | 0.001 | -0.005 | -0.001 |
| CRIC74SD | -0.005 | -0.23 | 0.004* | 0.005 | 0.002 | -0.009 | -0.002 |
| ADHD*CRIC15SD | 0.015 | 0.21 | 0.01 | 0.055 | 0.008 | -0.001 | 0.030 |
| ADHD*CRIC44SD | 0.017 | 0.20 | 0.014* | 0.029 | 0.008 | 0.000 | 0.030 |

| Hemodynamic response mean to go/no-go false alarms (FA): |  |  |  |  |  |  |  |
| --- | --- | --- | --- | --- | --- | --- | --- |
| Adj. $R^2$ = 0.28 | $df$ = 104 | | $F$ = 9.66 | | p <0.001 | | |
|  |  |  |  | Bootstrapping: |  |  |  |
| Predictors | B | $\beta$ | $p$ | $p$ | SE | $p$ 95% CI | |
| Age | -0.006 | -0.23 | 0.008* | 0.002 | 0.002 | -0.009 | -0.002 |
| Sex | -0.012 | -0.11 | 0.199 | 0.252 | 0.010 | -0.031 | 0.008 |
| WASI Full-scale IQ | 0.000 | -0.08 | 0.359 | 0.422 | 0.000 | -0.001 | 0.000 |
| ADHD | 0.034 | 0.27 | <0.001* | 0.001 | 0.008 | 0.018 | 0.048 |
| ADHD*CRIC15M | 0.018 | 0.28 | 0.001* | 0.005 | 0.006 | 0.006 | 0.031 |

| Hemodynamic response mean to go/no-go hits: |  |  |  |  |  |  |  |
| --- | --- | --- | --- | --- | --- | --- | --- |
| Adj. $R^2$ = 0.37 | $df$ = 102 | | $F$ = 10.25 | | p <0.001 | | |
|  |  |  |  | Bootstrapping: |  |  |  |
| Predictors | B | $\beta$ | $p$ | $p$ | SE | $p$ 95% CI | |
| Age | -0.008 | -0.30 | <0.001* | 0.001 | 0.002 | -0.011 | -0.004 |
| Sex | -0.004 | -0.03 | 0.67 | 0.690 | 0.010 | -0.023 | 0.014 |
| WASI Full-scale IQ | 1.232 | 0.00 | 0.97 | 0.969 | 0.000 | -0.001 | 0.001 |
| ADHD | 0.034 | 0.38 | <0.001* | 0.001 | 0.007 | 0.019 | 0.048 |
| Hits_IC18_M | -0.029 | -0.28 | <0.001* | 0.001 | 0.006 | -0.040 | -0.017 |
| ADHD*HitsIC37M | -0.019 | -0.21 | 0.008* | 0.014 | 0.010 | -0.042 | 0.001 |
| ADHD*HitsIC62M | -0.013 | -0.21 | 0.01* | 0.027 | 0.006 | -0.024 | 0.001 |

| Models^ with no predictions of hemodynamic responses on IIVRT: |  |  |  |  |  |  |  |
| --- | --- | --- | --- | --- | --- | --- | --- |
| Adj. $R^2$ = 0.21 | $df$ = 105 | | $F$ = 8.29 | | p <0.001 | | |
|  |  |  |  | Bootstrapping: |  |  |  |
| Predictors | B | $\beta$ | $p$ | $p$ | SE | 95% CI for B | |
| Age | -0.007 | -0.26 | 0.003* | 0.005* | 0.002 | -0.011 | -0.003 |
| Sex | -0.007 | -0.07 | 0.455 | 0.500 | 0.010 | -0.029 | 0.012 |
| WASI Full-scale IQ | 0.000 | -0.07 | 0.462 | 0.507 | 0.000 | -0.001 | 0.001 |
| ADHD | 0.033 | 0.37 | <0.001* | 0.001* | 0.008 | 0.017 | 0.050 |

Note. \* $p$ <0.017 + $p$ <0.05 in the bootstrapped resampling procedure; #= bootstrapped regression models; ^= regression models with no ICs predicting IIVRT= hemodynamic response variability to false alarms (FA) and hits, and further, hemodynamic response mean to correct rejections (CR); Adj. = adjusted; SE= standard error.
Note. \* $p$ <0.017 + $p$ <0.05 in the bootstrapped resampling procedure;

#### False Alarms and Hits

The results from the stepwise regression analyses of hemodynamic response variability to False Alarms and Hits as predictors, respectively, showed that only a diagnosis of ADHD and the covariate of lower age significantly predicted higher IIVRT, and not variability in hemodynamic responses, sex, or full-scale IQ (see right-hand columns of Table 2, Supplementary Table 4, and Supplementary Table 5).

### Mean amplitude of hemodynamic responses on level of IIVRT

#### Correct Rejections

The results from the bootstrapped regression model that predicted maximized variance showed no significant effects of mean hemodynamic responses on level of IIVRT. A diagnosis of ADHD and lower age did significantly predict higher IIVRT, whereas sex and full-scale IQ did not.

#### False Alarms

There was a significant interaction effect between amplitude mean to False Alarms in a bilateral superior temporal cortex with extensions to insula (IC 15) and a diagnosis of ADHD in predicting higher IIVRT. Higher amplitude variability in IC 15 specifically predicted higher IIVRT in ADHD. In addition, a diagnosis of ADHD and lower age predicted higher IIVRT as main effects, whereas sex and IQ did not.

#### Hits

The results from the bootstrapped regression model that predicted maximized variance showed significant interaction effects between amplitude means to Hits in IC 37 and IC 62, respectively, and ADHD on IIVRT. In addition, main effects appeared of mean amplitude in IC 18 and a diagnosis of ADHD, respectively, to predict higher IIVRT together with a lower age. Sex and IQ did not predict level of IIVRT.

## Discussion

The aim of the current study was to better localize and understand how brain activity variability might predict attentional lapses measured as high IIVRT in ADHD. As predicted, we found posterior DMN region variability in hemodynamic responses (IC 44 and IC 74), but not mean activity levels, predicted behavioral IIVRT on a Go/NoGo task. Higher variability in a PCC DMN (IC 44) predicted higher IIVRT only in ADHD. In contrast, variability in brain response for a precuneus DMN (IC 74) predicted lower IIVRT both in ADHD and healthy controls. Further, mean amplitude of hemodynamic responses in a bilateral superior temporal network with extensions to insula (IC 15) specifically predicted higher IIVRT in ADHD and not healthy controls. These findings appear to be the first that show posterior DMN is involved in fluctuation of attention in ADHD. This can be viewed as important support for a DMN hypothesis (16) since the posterior DMN node was originally showed to have the highest spontaneous fluctuations during resting state (13, 26). We did not, however, as expected find an effect of moment-to-moment fluctuations in the anterior DMN regions or an ADHD-specific effect of mean hemodynamic responses in right motor systems on level of attentional lapses. Importantly, the findings provide another clear indication that the consistency with which specific brain regions – especially the DMN – engage from trial-to-trial are particularly important to understanding the neurobiology of attentional lapses in ADHD.

Variability of hemodynamic responses elicited by correctly rejected NoGo stimuli played different roles in ADHD for different posterior DMN brain regions. For the PCC (IC 44), greater inconsistency of activation predicted higher IIVRT for ADHD adolescents, but in the precuneus (IC 74) more inconsistent responses led to lower IIVRT. In many fMRI studies, the PCC and precuneus comprise one DMN subnetwork (54). The precuneus is recognized to be a functional core hub in connection with other functional networks that has an important role monitoring and regulating attention fluctuations (25), possibly rapidly adjusting attention resources in response to contextual changes. The schism in ADHD between brain variability versus behavioral variability for these two regions that normally closely co-engage across different task contexts emphasizes their dysfunction in ADHD. Thus, higher variability in hemodynamic responses in ADHD may not only be a potential marker of poorer attention functioning, but might provide a clue that important intra-network dysfunction or disconnection might be a specific factor that gives rise to abnormal IIVRT. This underscores the importance of studying study-elicited brain function; resting-state fMRI studies can only show high/low DMN activity fluctuations compared to control groups, whereas task-based studies can additionally quantify the consistency of that activity in different task-relevant contexts.

Interestingly, similar to previous fMRI studies using Go/NoGo tasks (30-32), there was no evidence in this study that anterior DMN areas were involved in IIVRT. In contrast, such evidence has been found in other fMRI studies, but it is not currently clear if the task choice (e.g., oddball (23, 24) versus working memory (22)) or the fact that some of those tasks were self-paced might be responsible for anterior DMN involvement in IIVRT. We speculate that speeded reaction time tasks requiring a higher load on sustained attention compared to self-paced tasks might involve posterior DMN due to requirements of sustaining voluntary-controlled neural brain activation over time. Regardless of the ultimate explanation, more studies should examine the neural correlates of IIVRT using a variety of different paradigms for a complete understanding.

We also found specifically for ADHD-diagnosed adolescents that higher IIVRT was predicted by mean hemodynamic responses to failed NoGo responses (false alarms) in a bilateral superior temporal network with extensions medially to insula (IC 15). To our knowledge, three prior studies linked the average levels of hemodynamic responses in temporal lobes to how much IIVRT is observed in ADHD (23, 32, 55). In one study of a small sample of ADHD boys (23), higher mean hemodynamic responses in a superior temporal cortex network with extensions to insula was associated with lower IIVRT on a visual oddball task, similarly to what we found in this study in healthy controls. The superior temporal lobes are mostly known for the involvement in language processing and as the location of the primary auditory cortex. However, several studies have shown that this area also is important for attention allocation (56), particularly involuntary orienting attention (57). Interestingly, the Rubia et al. (23) study showed that lower mean hemodynamic responses bilaterally in superior and medial temporal cortices to a visual oddball stimuli that require attention orientation (57) appeared only in the ADHD group.

Furthermore, the insula (part of IC 15), is an important salience network node involved in alerting/arousal attention, which typically interacts with involuntary orienting attention (58). Collectively, these observations raise the possibility the adolescents with ADHD in our study might have been distracted by external/internal salient stimuli when they were to focus on inhibiting motor responses. Alternatively, this IIVRT-temporal lobe relationship might reflect brain activity related to some sort of compensatory mechanism to help direct attention, secondary to the posterior DMN abnormalities we observed in the ADHD group.

Both higher mean and variability of hemodynamic responses across the Go/NoGo parameters in a right hemisphere postcentral gyrus motor system (IC 18) predicted higher IIVRT both in the adolescents with ADHD and healthy controls. Further, mean amplitudes to hits in IC 37 and IC 62, respectively, predicted higher IIVRT specifically in ADHD and not healthy controls. Important to note is that the Go/NoGo task used in the current study included a jittering design by varying inter-stimulus-intervals. The jittering design can introduce challenges in reliably isolating hemodynamic response amplitude or variability to the Go responses (Hits) across trials (59). Compounding this, the ‘X’ stimuli were frequent and rapid enough to nearly saturate the hemodynamic response to this condition. As such, Hit-related findings should perhaps be interpreted conservatively until they have been replicated.

Study strengths included the moderate-sized sample to lend confidence in the results, the use of data-reduction ICA methodology to concurrently consider all major brain network regions in the same analysis in a statistically efficient way, and use of data-driven hypothesis-testing to remain unconstrained by rigid theoretical assumptions of the neurobiological underpinnings of high IIVRT in ADHD. In contrast, because the sample size was not definitive, the results of this Go/NoGo fMRI study of IIVRT in ADHD require replication.

## Conclusion

Our findings suggest that variability in hemodynamic responses in posterior DMN elicited on rapid tests with high cognitive demands are important correlates of higher IIVRT during Go/NoGo task performance for ADHD-diagnosed adolescents. Higher behavioral IIVRT is a recognized marker of poorer brain health (60) and predicts all-cause mortality in adults (61).

Further, moment-to-moment fluctuations of brain signaling also has been suggested as an important parameter of brain health (36, 52). These facts not only emphasizes the general importance of better understanding the neural mechanisms that give rise to IIVRT, they underscores the need to characterize how dysfunction in this neurobiology yields the typically high IIVRT found in ADHD-diagnosed patients. Such a mechanistic understanding could provide important clues to disorder pathophysiology, as there already is evidence that higher amplitude variability in resting-state fMRI (Nomi et al., 2018) and during fMRI attention processing (55) predict higher ADHD symptom severity. As such, IIVRT-related neural dysfunction in ADHD holds potential to be treatment target for the disorder.

## Supporting information

https://www.researchgate.net/publication/358365077_Moment-to-moment_fluctuations_of_hemodynamic_responses_in_posterior_default_mode_networks_different

## Data Availability

All data produced in the present study are available upon reasonable request to the authors

## Acknowledgements

This project was funded by National Institutes of Health Grant No. R01MH080956 (to MCS). We thank the adolescents and their parents for participating in the study and the research staff on the project, including Danielle Francois, Nicole Pompay, Ethan Rosenfeld, and Christina Wong. We would also thank Dr. Tom Eichele for sharing his script for ICA single-trial estimates of hemodynamic responses.

The submitted version of this manuscript has been published as a preprint:

Sørensen, L., Chung, Y.S., Khadka, S., & Stevens, M.C. (2022). Moment-to-moment fluctuations of hemodynamic responses in posterior default mode networks differentially predicts level of attentional lapses in adolescents with Attention-Deficit/Hyperactivity Disorder. *medRxiv*. https://doi.org/10.1101/2022.01.31.22270169

## References

1. Polanczyk G, Jensen P (2008): Epidemiologic considerations in attention deficit hyperactivity disorder: a review and update. Child Adolesc Psychiatr Clin N Am. 17:245–260, vii.

2. Castellanos FX, Tannock R (2002): Neuroscience of attention-deficit/hyperactivity disorder: the search for endophenotypes. Nature reviews Neuroscience. 3:617–628.

3. Kofler MJ, Rapport MD, Sarver DE, Raiker JS, Orban SA, Friedman LM, et al. (2013): Reaction time variability in ADHD: a meta-analytic review of 319 studies. Clinical psychology review. 33:795–811.

4. Kuntsi J, Klein C (2012): Intraindividual variability in ADHD and its implications for research of causal links. Current topics in behavioral neurosciences. 9:67–91.

5. Tamm L, Narad ME, Antonini TN, O’Brien KM, Hawk LW, Jr., Epstein JN (2012): Reaction time variability in ADHD: a review. Neurotherapeutics : the journal of the American Society for Experimental NeuroTherapeutics. 9:500–508.

6. Weissman DH, Roberts KC, Visscher KM, Woldorff MG (2006): The neural bases of momentary lapses in attention. Nature neuroscience. 9:971–978.

7. Epstein JN, Langberg JM, Rosen PJ, Graham A, Narad ME, Antonini TN, et al. (2011): Evidence for higher reaction time variability for children with ADHD on a range of cognitive tasks including reward and event rate manipulations. Neuropsychology. 25:427–441.

8. Karalunas SL, Geurts HM, Konrad K, Bender S, Nigg JT (2014): Annual research review: Reaction time variability in ADHD and autism spectrum disorders: measurement and mechanisms of a proposed trans-diagnostic phenotype. Journal of child psychology and psychiatry, and allied disciplines. 55:685–710.

9. Bellgrove MA, Hawi Z, Kirley A, Gill M, Robertson IH (2005): Dissecting the attention deficit hyperactivity disorder (ADHD) phenotype: sustained attention, response variability and spatial attentional asymmetries in relation to dopamine transporter (DAT1) genotype. Neuropsychologia. 43:1847–1857.

10. Kuntsi J, Oosterlaan J, Stevenson J (2001): Psychological mechanisms in hyperactivity: I. Response inhibition deficit, working memory impairment, delay aversion, or something else? Journal of child psychology and psychiatry, and allied disciplines. 42:199–210.

11. Antonini TN, Narad ME, Langberg JM, Epstein JN (2013): Behavioral correlates of reaction time variability in children with and without ADHD. Neuropsychology. 27:201–209.

12. Adamo N, Hodsoll J, Asherson P, Buitelaar JK, Kuntsi J (2019): Ex-Gaussian, Frequency and Reward Analyses Reveal Specificity of Reaction Time Fluctuations to ADHD and Not Autism Traits. J Abnorm Child Psychol. 47:557–567.

13. Raichle ME, MacLeod AM, Snyder AZ, Powers WJ, Gusnard DA, Shulman GL (2001): A default mode of brain function. Proceedings of the National Academy of Sciences of the United States of America. 98:676–682.

14. Shulman GL, Fiez JA, Corbetta M, Buckner RL, Miezin FM, Raichle ME, et al. (1997): Common Blood Flow Changes across Visual Tasks: II. Decreases in Cerebral Cortex. J Cogn Neurosci. 9:648–663.

15. Castellanos FX, Kelly C, Milham MP (2009): The restless brain: attention-deficit hyperactivity disorder, resting-state functional connectivity, and intrasubject variability. Can J Psychiatry. 54:665–672.

16. Sonuga-Barke EJ, Castellanos FX (2007): Spontaneous attentional fluctuations in impaired states and pathological conditions: a neurobiological hypothesis. Neuroscience and biobehavioral reviews. 31:977–986.

17. Castellanos FX, Margulies DS, Kelly C, Uddin LQ, Ghaffari M, Kirsch A, et al. (2008): Cingulate-precuneus interactions: a new locus of dysfunction in adult attention-deficit/hyperactivity disorder. Biological psychiatry. 63:332–337.

18. Choi J, Jeong B, Lee SW, Go HJ (2013): Aberrant development of functional connectivity among resting state-related functional networks in medication-naive ADHD children. PLoS One. 8:e83516.

19. Fair DA, Posner J, Nagel BJ, Bathula D, Dias TG, Mills KL, et al. (2010): Atypical default network connectivity in youth with attention-deficit/hyperactivity disorder. Biological psychiatry. 68:1084–1091.

20. Sripada C, Kessler D, Fang Y, Welsh RC, Prem Kumar K, Angstadt M (2014): Disrupted network architecture of the resting brain in attention-deficit/hyperactivity disorder. Hum Brain Mapp. 35:4693–4705.

21. Sun L, Cao Q, Long X, Sui M, Cao X, Zhu C, et al. (2012): Abnormal functional connectivity between the anterior cingulate and the default mode network in drug-naive boys with attention deficit hyperactivity disorder. Psychiatry Res. 201:120–127.

22. Fassbender C, Zhang H, Buzy WM, Cortes CR, Mizuiri D, Beckett L, et al. (2009): A lack of default network suppression is linked to increased distractibility in ADHD. Brain research. 1273:114–128.

23. Rubia K, Smith AB, Brammer MJ, Taylor E (2007): Temporal lobe dysfunction in medication-naive boys with attention-deficit/hyperactivity disorder during attention allocation and its relation to response variability. Biological psychiatry. 62:999–1006.

24. Sorensen L, Eichele T, van Wageningen H, Plessen KJ, Stevens MC (2016): Amplitude variability over trials in hemodynamic responses in adolescents with ADHD: The role of the anterior default mode network and the non-specific role of the striatum. Neuroimage Clin. 12:397–404.

25. Cavanna AE, Trimble MR (2006): The precuneus: a review of its functional anatomy and behavioural correlates. Brain. 129:564–583.

26. Greicius MD, Krasnow B, Reiss AL, Menon V (2003): Functional connectivity in the resting brain: a network analysis of the default mode hypothesis. Proceedings of the National Academy of Sciences of the United States of America. 100:253–258.

27. Cortese S, Kelly C, Chabernaud C, Proal E, Di Martino A, Milham MP, et al. (2012): Toward systems neuroscience of ADHD: a meta-analysis of 55 fMRI studies. Am J Psychiatry. 169:1038–1055.

28. Hart H, Radua J, Nakao T, Mataix-Cols D, Rubia K (2013): Meta-analysis of functional magnetic resonance imaging studies of inhibition and attention in attention-deficit/hyperactivity disorder: exploring task-specific, stimulant medication, and age effects. JAMA Psychiatry. 70:185–198.

29. Lei D, Du M, Wu M, Chen T, Huang X, Du X, et al. (2015): Functional MRI reveals different response inhibition between adults and children with ADHD. Neuropsychology. 29:874–881.

30. Siniatchkin M, Glatthaar N, von Muller GG, Prehn-Kristensen A, Wolff S, Knochel S, et al. (2012): Behavioural treatment increases activity in the cognitive neuronal networks in children with attention deficit/hyperactivity disorder. Brain Topogr. 25:332–344.

31. Suskauer SJ, Simmonds DJ, Caffo BS, Denckla MB, Pekar JJ, Mostofsky SH (2008): fMRI of intrasubject variability in ADHD: anomalous premotor activity with prefrontal compensation. Journal of the American Academy of Child and Adolescent Psychiatry. 47:1141–1150.

32. Spinelli S, Vasa RA, Joel S, Nelson TE, Pekar JJ, Mostofsky SH (2011): Variability in post-error behavioral adjustment is associated with functional abnormalities in the temporal cortex in children with ADHD. Journal of child psychology and psychiatry, and allied disciplines. 52:808–816.

33. van Belle J, van Raalten T, Bos DJ, Zandbelt BB, Oranje B, Durston S (2015): Capturing the dynamics of response variability in the brain in ADHD. Neuroimage Clin. 7:132–141.

34. Mostofsky SH, Simmonds DJ (2008): Response inhibition and response selection: two sides of the same coin. J Cogn Neurosci. 20:751–761.

35. Stevens MC, Pearlson GD, Calhoun VD, Bessette KL (2018): Functional Neuroimaging Evidence for Distinct Neurobiological Pathways in Attention-Deficit/Hyperactivity Disorder. Biol Psychiatry Cogn Neurosci Neuroimaging. 3:675–685.

36. Boylan MA, Foster CM, Pongpipat EE, Webb CE, Rodrigue KM, Kennedy KM (2021): Greater BOLD Variability is Associated With Poorer Cognitive Function in an Adult Lifespan Sample. Cereb Cortex. 31:562–574.

37. Eichele T, Debener S, Calhoun VD, Specht K, Engel AK, Hugdahl K, et al. (2008): Prediction of human errors by maladaptive changes in event-related brain networks. Proceedings of the National Academy of Sciences of the United States of America. 105:6173–6178.

38. Suskauer SJ, Simmonds DJ, Fotedar S, Blankner JG, Pekar JJ, Denckla MB, et al. (2008): Functional magnetic resonance imaging evidence for abnormalities in response selection in attention deficit hyperactivity disorder: differences in activation associated with response inhibition but not habitual motor response. J Cogn Neurosci. 20:478–493.

39. Kaufman J, Birmaher B, Brent D, Rao U, Flynn C, Moreci P, et al. (1997): Schedule for Affective Disorders and Schizophrenia for School-Age Children-Present and Lifetime Version (K-SADS-PL): initial reliability and validity data. Journal of the American Academy of Child and Adolescent Psychiatry. 36:980–988.

40. Kadesjo B, Gillberg C (2001): The comorbidity of ADHD in the general population of Swedish school-age children. Journal of child psychology and psychiatry, and allied disciplines. 42:487–492.

41. Wechsler D (1999): Wechsler Abbrievated Scale of Intelligence. WASI. Manual. San Antonio, Tex: The Psychological Corporation.

42. Stevens MC, Kiehl KA, Pearlson GD, Calhoun VD (2007): Functional neural networks underlying response inhibition in adolescents and adults. Behav Brain Res. 181:12–22.

43. Jenkinson M, Bannister P, Brady M, Smith S (2002): Improved optimization for the robust and accurate linear registration and motion correction of brain images. Neuroimage. 17:825–841.

44. Jenkinson M, Beckmann CF, Behrens TE, Woolrich MW, Smith SM (2012): Fsl. Neuroimage. 62:782–790.

45. Andersson JL, Hutton C, Ashburner J, Turner R, Friston K (2001): Modeling geometric deformations in EPI time series. Neuroimage. 13:903–919.

46. Cox RW (1996): AFNI: software for analysis and visualization of functional magnetic resonance neuroimages. Comput Biomed Res. 29:162–173.

47. Griffanti L, Salimi-Khorshidi G, Beckmann CF, Auerbach EJ, Douaud G, Sexton CE, et al. (2014): ICA-based artefact removal and accelerated fMRI acquisition for improved resting state network imaging. Neuroimage. 95:232–247.

48. Abou-Elseoud A, Starck T, Remes J, Nikkinen J, Tervonen O, Kiviniemi V (2010): The effect of model order selection in group PICA. Human brain mapping. 31:1207–1216.

49. Kiviniemi V, Starck T, Remes J, Long X, Nikkinen J, Haapea M, et al. (2009): Functional segmentation of the brain cortex using high model order group PICA. Human brain mapping. 30:3865–3886.

50. Bell AJ, Sejnowski TJ (1995): An information-maximization approach to blind separation and blind deconvolution. Neural computation. 7:1129–1159.

51. Erhardt EB, Rachakonda S, Bedrick EJ, Allen EA, Adali T, Calhoun VD (2011): Comparison of multi-subject ICA methods for analysis of fMRI data. Human brain mapping. 32:2075–2095.

52. Mohr PN, Nagel IE (2010): Variability in brain activity as an individual difference measure in neuroscience? J Neurosci. 30:7755–7757.

53. Leys C, Lay C, Klein O, Bernard P, Licata L (2013): Detecting outliers: do not use standard deviation around the mean, use absolute deviation aroudn the median. Journal of Experimental Social Psychology. 49:764–766.

54. Wang S, Tepfer LJ, Taren AA, Smith DV (2020): Functional parcellation of the default mode network: a large-scale meta-analysis. Sci Rep. 10:16096.

55. Depue BE, Burgess GC, Willcutt EG, Bidwell LC, Ruzic L, Banich MT (2010): Symptom-correlated brain regions in young adults with combined-type ADHD: their organization, variability, and relation to behavioral performance. Psychiatry Res. 182:96–102.

56. Kobel M, Bechtel N, Specht K, Klarhofer M, Weber P, Scheffler K, et al. (2010): Structural and functional imaging approaches in attention deficit/hyperactivity disorder: does the temporal lobe play a key role? Psychiatry Res. 183:230–236.

57. Kim H (2014): Involvement of the dorsal and ventral attention networks in oddball stimulus processing: a meta-analysis. Hum Brain Mapp. 35:2265–2284.

58. Posner MI (2012): Attentional networks and consciousness. Front Psychol. 3:64.

59. Amaro E, Jr., Barker GJ (2006): Study design in fMRI: basic principles. Brain Cogn. 60:220–232.

60. MacDonald SW, Nyberg L, Backman L (2006): Intra-individual variability in behavior: links to brain structure, neurotransmission and neuronal activity. Trends Neurosci. 29:474–480.

61. Shipley BA, Der G, Taylor MD, Deary IJ (2006): Cognition and all-cause mortality across the entire adult age range: health and lifestyle survey. Psychosom Med. 68:17–24.

