## Supplementary material for "Moment-to-moment fluctuations of hemodynamic responses in posterior default mode networks differentially predict level of attentional lapses in adolescents with Attention-Deficit/Hyperactivity Disorder": https://www.researchgate.net/publication/358365077_Moment-to-moment_fluctuations_of_hemodynamic_responses_in_posterior_default_mode_networks_different

Supplemental Table 1. The ICs that did not predict level of IIVRT in relation to variability in hemodynamic responses.

| ICs | Brain networks | Max Peak | | Figures |
|---|---|---|---|---|
| | | *x* , *y* , *z* | *t* | |
| IC 1 | Bilateral Precentral Gyrus | -50, -12, 34 | 34.26 | 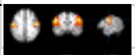 |
| IC 4 | Right Superior Temporal Gyrus | 32, 8, -22 | 35.46 | 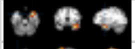 |
| IC 6 | Left precentral/postcentral Gyrus | -34, -24, 52 | 24.23 | 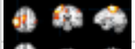 |
| IC 13 | Bilateral Thalamus | -12, -6, 16 | 26.08 | 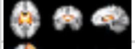 |
| IC 14 | Right Middle Frontal Gyrus | 44, 48, 22 | 23.06 | 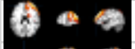 |
| IC 17 | Bilateral superior parietal/Precuneus | -4, -48, 60 | 18.88 | 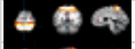 |
| IC 21 | Right Middle Temporal Gyrus | 56, -32, -2 | 20.20 | 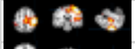 |
| IC 23 | Left Parahippocampus | -22, 0, -14 | 23.77 | 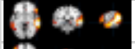 |
| IC 28 | Left Inferior Frontal Gyrus | -52, 18, 4 | 24.27 | 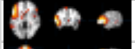 |
| IC 31 | Right Medial Frontal Gyrus | 4, 50, 38 | 20.08 | 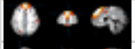 |
| IC 33 | Bilateral Superior Parietal Cortex | -22, -66, 46 | 16.05 | 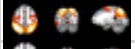 |
| IC 35 | Bilateral Posterior Cingulate Cortex* | -2, -44, 8 | 22.64 | 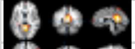 |
| IC 36 | Bilateral Putamen | -16, 4, -8 | 18.57 | 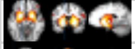 |
| IC 37 | Bilateral Parietal Postcentral Gyrus | -52,-26, 40 | 19.59 | 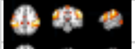 |
| IC 39 | Bilateral Cingulate Gyrus* | -6, -34, 26 | 21.73 | 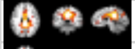 |
| IC 40 | Left Parietal Angular Gyrus | -50, -62, 34 | 21.17 | 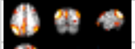 |
| IC 41 | Right Inferior Frontal Gyrus | 50, 40, -2 | 23.75 | 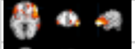 |
| IC 42 | Right Inferior Parietal Lobe | 54, -50, 42 | 16.38 | 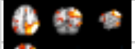 |
| IC 46 | Right Middle Frontal Gyrus | 46, 40, 18 | 16.42 | 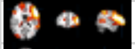 |
| IC 47 | Left Rostral Middle Frontal Gyrus | -50, 8, 40 | 19.33 | 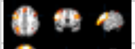 |
| IC 50 | Bilateral Rostral Anterior Cingulate Cortex* | -4, 38, 14 | 19.25 | 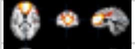 |
| IC 53 | Bilateral Precuenus* | 4, -74, 40 | 24.19 | 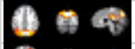 |
| IC 54 | Left Middle Temporal Gyrus | -58, -30, -6 | 19.67 | 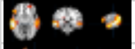 |
| IC 55 | Bilateral Lateral Orbitofrontal Cortex | 40, 34, -18 | 21.66 | 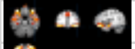 |
| IC 62 | Bilateral Rostral Middle Frontal Gyrus | 28, 60, 6 | 19.63 | 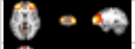 |
| IC 64 | Bilateral Inferior Parietal Cortex | 38, -74, 32 | 17.30 | 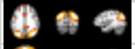 |
| IC 70 | Bilateral Medial Anterior Cingulate Cortex* | -4, 52, -2 | 21.79 | 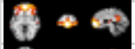 |
| IC 71 | Bilateral Precuenus* | 4, -50, 30 | 17.80 | 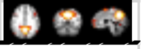 |

es. Note. *= Default Mode Networks. IC = Indenpendent Component.

Supplemental Table 2. Bivariate correlations between the descriptive sample variables.

| Variables: | Mean RT | Age | Sex | Full-scale IQ |
|---|---|---|---|---|
| IIVRT | 0.33** | -0.29** | -0.11 | -0.16 |
| Mean RT | | -0.13 | -0.25** | -0.11 |
| Age | | | 0.14 | 0.03 |
| Sex | | | | 0.08 |
| Full-scale IQ | | | | |

Note. ***p* <0.01; **p* <.05; IIVRT= Intraindividual Variability in Reaction Time; RT= Reaction Times; ADHD= Attention Deficit/ Hyperactivity Disorder.

Supplemental Table 3. Bivariate correlations between variability in hemodynamic responses to Correct Rejections and IIVRT.

| | Amplitude variability in hemodynamic responses to go/no-go correct rejections (CR): | | | | | | | | | | | | | | | | | | | | | | | | | | | | | | Behavioral variables | | | | | |
|---|---|---|---|---|---|---|---|---|---|---|---|---|---|---|---|---|---|---|---|---|---|---|---|---|---|---|---|---|---|---|---|---|---|---|---|---|
| ICs | IC 4 | IC 6 | IC 13 | IC 14 | IC 15 | IC 17 | IC 18 | IC 21 | IC 23 | IC 28 | IC 31 | IC 33 | IC 35 | IC 36 | IC 37 | IC 39 | IC 40 | IC 41 | IC 42 | IC 44 | IC 46 | IC 47 | IC 50 | IC 53 | IC 54 | IC 55 | IC 62 | IC 64 | IC 70 | IC 71 | IC 74 | IIVRT | Mean RT | Age | ADHD | Sex | FSIQ |
| IC 1 | -0.08 | 0.27** | 0.42** | 0.28** | 0.35** | 0.33** | 0.30** | 0.26** | 0.27** | 0.27** | 0.21* | 0.32** | 0.07 | 0.33** | 0.34** | 0.25** | 0.23* | 0.29** | 0.30** | 0.00 | 0.36** | 0.18 | 0.27** | 0.30** | 0.26** | 0.15 | 0.26** | 0.23* | 0.25** | 0.10 | 0.24* | -0.15 | -0.03 | 0.02 | -0.04 | 0.11 | -0.01 |
| IC 4 | | 0.11 | -0.05 | 0.19* | -0.08 | -0.01 | 0.10 | 0.14 | 0.09 | 0.05 | 0.06 | 0.05 | -0.04 | 0.13 | 0.02 | 0.07 | -0.11 | 0.08 | 0.07 | 0.10 | 0.18 | 0.09 | 0.01 | -0.02 | -0.02 | 0.25** | 0.17 | -0.07 | 0.08 | 0.06 | 0.04 | -0.15 | 0.00 | 0.04 | 0.01 | 0.03 | 0.07 |
| IC 6 | | | 0.31** | 0.28** | 0.27** | 0.35** | 0.50** | 0.27** | 0.30** | 0.19* | 0.02 | 0.15 | 0.18 | 0.22* | 0.27** | 0.18 | 0.36** | 0.07 | 0.35** | 0.19* | 0.17 | 0.22* | 0.15 | 0.40** | 0.20* | 0.24* | 0.28** | 0.02 | 0.03 | 0.06 | 0.08 | -0.12 | -0.14 | 0.05 | -0.15 | 0.08 | 0.03 |
| IC 13 | | | | 0.04 | 0.35** | 0.19* | 0.15 | 0.28** | 0.37** | 0.36** | 0.10 | 0.11 | 0.09 | 0.18 | 0.14 | 0.13 | 0.21* | 0.27** | 0.07 | 0.14 | 0.17 | 0.12 | -0.00 | 0.38** | 0.02 | 0.20* | 0.23* | 0.16 | 0.18 | 0.06 | 0.29** | -0.10 | 0.01 | -0.04 | -0.09 | -0.01 | -0.06 |
| IC 14 | | | | | 0.21* | 0.20* | 0.28** | 0.21* | 0.20* | 0.23* | 0.19* | 0.15 | -0.05 | 0.15 | 0.14 | 0.15 | 0.10 | 0.11 | 0.14 | 0.13 | 0.13 | 0.31** | 0.24* | 0.21* | 0.19 | 0.33** | 0.17 | -0.03 | 0.11 | 0.08 | 0.01 | -0.09 | 0.05 | -0.00 | -0.09 | -0.01 | 0.04 |
| IC 15 | | | | | | 0.31** | 0.37** | 0.32** | 0.31** | 0.26** | 0.19* | 0.24* | 0.17 | 0.27** | 0.24* | 0.35** | 0.32** | 0.27** | 0.14 | 0.33** | 0.34** | 0.32** | 0.27** | 0.40** | 0.27** | 0.20* | 0.38** | 0.16 | 0.28** | 0.08 | 0.31** | 0.03 | -0.09 | -0.13 | -0.06 | 0.15 | 0.18 |
| IC 17 | | | | | | | 0.20* | 0.23* | 0.17 | 0.19* | 0.14 | 0.33** | 0.13 | 0.14 | 0.11 | 0.23* | 0.30** | 0.16 | 0.33** | 0.32** | 0.21* | 0.15 | 0.23* | 0.40** | 0.06 | 0.12 | 0.16 | 0.18 | 0.20* | -0.05 | 0.15 | -0.02 | 0.03 | 0.14 | 0.04 | 0.03 | 0.14 |
| IC 18 | | | | | | | | 0.12 | 0.25** | 0.19* | 0.07 | 0.36** | -0.03 | 0.24* | 0.22* | 0.29** | 0.23* | 0.15 | 0.34** | 0.10 | 0.29** | 0.25** | 0.08 | 0.29** | 0.32** | 0.26** | 0.19* | 0.11 | 0.20* | 0.19 | 0.14 | -0.24* | -0.24* | -0.10 | -0.09 | 0.13 | 0.04 |
| IC 21 | | | | | | | | | 0.33** | 0.22* | 0.22* | 0.17 | 0.16 | 0.23* | 0.19* | 0.28** | 0.24* | 0.29** | 0.17 | 0.13 | 0.27** | 0.21* | 0.27** | 0.28** | 0.17 | 0.18 | 0.33** | 0.10 | 0.10 | 0.10 | 0.16 | -0.06 | 0.01 | 0.02 | -0.07 | 0.01 | 0.07 |
| IC 23 | | | | | | | | | | 0.22* | 0.23* | 0.15 | 0.11 | 0.44** | 0.13 | 0.21* | 0.11 | 0.18 | 0.18 | 0.21* | 0.24* | 0.25** | -0.02 | 0.38** | 0.07 | 0.22* | 0.28** | 0.27** | 0.19 | 0.13 | 0.23* | -0.15 | -0.14 | 0.10 | -0.05 | -0.07 | 0.12 |
| IC 28 | | | | | | | | | | | 0.12 | 0.16 | -0.05 | 0.13 | 0.13 | 0.20* | 0.27** | 0.39** | 0.12 | 0.29** | 0.28** | 0.26** | 0.20* | 0.25** | 0.24* | 0.35** | 0.21* | 0.38** | 0.28** | 0.27** | 0.40** | -0.15 | -0.09 | 0.15 | -0.04 | 0.17 | 0.05 |
| IC 31 | | | | | | | | | | | | 0.19* | -0.18 | 0.11 | 0.00 | 0.08 | -0.07 | 0.05 | 0.14 | 0.12 | -0.07 | 0.07 | 0.16 | 0.01 | -0.04 | 0.05 | 0.22* | 0.05 | 0.20* | -0.01 | 0.18 | -0.02 | -0.05 | 0.22* | 0.11 | 0.06 | 0.00 |
| IC 33 | | | | | | | | | | | | | -0.00 | 0.09 | 0.17 | 0.23* | 0.38** | 0.19* | 0.46** | 0.23* | 0.32** | 0.06 | 0.10 | 0.33** | 0.11 | 0.05 | 0.18 | 0.28** | 0.34** | 0.15 | 0.19* | -0.13 | -0.13 | 0.04 | -0.01 | 0.03 | 0.10 |
| IC 35 | | | | | | | | | | | | | | 0.10 | 0.16 | 0.06 | 0.05 | -0.10 | 0.02 | 0.03 | 0.08 | 0.05 | 0.06 | 0.06 | -0.10 | 0.05 | 0.22* | -0.06 | -0.06 | -0.02 | -0.01 | -0.02 | -0.08 | 0.10 | 0.09 | 0.18 | 0.05 |
| IC 36 | | | | | | | | | | | | | | | 0.27** | 0.30** | 0.09 | 0.12 | 0.10 | 0.01 | 0.22* | 0.12 | 0.10 | 0.27** | -0.00 | 0.00 | 0.21* | 0.17 | 0.11 | 0.32** | 0.23* | 0.06 | 0.12 | -0.04 | 0.11 | 0.07 | 0.09 |
| IC 37 | | | | | | | | | | | | | | | | 0.23* | 0.40** | 0.06 | 0.28** | 0.05 | 0.42** | 0.23* | 0.11 | 0.33** | 0.17 | -0.04 | 0.31** | 0.17 | 0.15 | 0.11 | 0.18 | 0.02 | -0.02 | -0.18 | -0.10 | 0.03 | -0.05 |
| IC 39 | | | | | | | | | | | | | | | | | 0.18 | 0.14 | 0.14 | 0.26** | 0.33** | 0.30** | 0.35** | 0.37** | 0.07 | 0.22* | 0.26** | 0.26** | 0.20* | 0.14 | 0.20* | -0.15 | -0.12 | -0.15 | -0.09 | 0.17 | 0.19* |
| IC 40 | | | | | | | | | | | | | | | | | | 0.13 | 0.34* | 0.29** | 0.39** | 0.33** | 0.15 | 0.53** | 0.21* | 0.08 | 0.33** | 0.32** | 0.25** | 0.22* | 0.22* | -0.06 | -0.04 | 0.08 | -0.12 | 0.16 | 0.10 |
| IC 41 | | | | | | | | | | | | | | | | | | | 0 .05 | 0.01 | 0.23* | 0.09 | 0.15 | 0.24* | 0.28** | 0.31** | 0.13 | 0.27** | 0.24* | 0.04 | 0.16 | -0.11 | 0.01 | -0.01 | 0.01 | -0.08 | -0.07 |
| IC 42 | | | | | | | | | | | | | | | | | | | | 0.23* | 0.29** | 0.09 | 0.12 | 0.35** | 0.11 | -0.05 | 0.09 | 0.19* | 0.22* | 0.20* | 0.09 | -0.06 | -0.06 | 0.08 | -0.07 | 0.03 | -0.12 |
| IC 44 | | | | | | | | | | | | | | | | | | | | | 0.15 | 0.19 | 0.16 | 0.37** | 0.12 | 0.05 | 0.10 | 0.187* | 0.17 | 0.07 | 0.24* | 0.07 | -0.17 | 0.07 | -0.03 | 0.16 | 0.07 |
| IC 46 | | | | | | | | | | | | | | | | | | | | | | 0.43** | 0.19 | 0.37** | 0.25** | 0.12 | 0.28** | 0.29** | 0.18 | 0.27** | 0.30** | -0.16 | -0.13 | -0.09 | -0.09 | 0.22* | 0.14 |
| IC 47 | | | | | | | | | | | | | | | | | | | | | | | 0.27** | 0.22* | 0.26** | 0.21* | 0.35** | 0.18 | 0.34** | 0.27** | 0.13 | -0.02 | -0.09 | 0.09 | 0.02 | 0.20* | 0.05 |
| IC 50 | | | | | | | | | | | | | | | | | | | | | | | | 0.24* | 0.21* | 0.19* | 0.28** | 0.08 | 0.29** | 0.15 | 0.14 | -0.05 | -0.05 | 0.10 | -0.01 | 0.29** | 0.02 |
| IC 53 | | | | | | | | | | | | | | | | | | | | | | | | | 0.18 | 0.18 | 0.28** | 0.35** | 0.32** | 0.16 | 0.22* | -0.08 | -0.05 | 0.06 | -0.04 | 0.11 | -0.02 |
| IC 54 | | | | | | | | | | | | | | | | | | | | | | | | | | 0.22* | 0.21* | 0.09 | 0.20* | 0.11 | 0.12 | -0.04 | -0.18 | -0.03 | -0.08 | 0.05 | 0.01 |
| IC 55 | | | | | | | | | | | | | | | | | | | | | | | | | | | 0.27** | 0.19* | 0.24* | 0.09 | 0.13 | -0.10 | 0.06 | 0.00 | -0.06 | 0.08 | 0.08 |
| IC 62 | | | | | | | | | | | | | | | | | | | | | | | | | | | | 0.05 | 0.23* | 0.08 | 0.17 | -0.02 | 0.06 | 0.11 | -0.02 | 0.05 | 0.15 |
| IC 64 | | | | | | | | | | | | | | | | | | | | | | | | | | | | | 0.34** | 0.24* | 0.40** | -0.14 | -0.32** | 0.08 | -0.07 | 0.12 | -0.03 |
| IC 70 | | | | | | | | | | | | | | | | | | | | | | | | | | | | | | 0.24* | 0.18 | -0.05 | -0.06 | 0.13 | 0.12 | 0.04 | -0.09 |
| IC 71 | | | | | | | | | | | | | | | | | | | | | | | | | | | | | | | .020* | -0.10 | -0.04 | 0.11 | 0.05 | 0.02 | 0.01 |
| IC 74 | | | | | | | | | | | | | | | | | | | | | | | | | | | | | | | | -0.23* | -0.07 | -0.03 | -0.15 | 0.11 | 0.15 |

Note. ***p*<.01; **p*<.05. IC = Indenpendent Component; IIVRT = Intraindividual Variability in Reaction Times; RT = Reaction Times; ADHD; Attention Deficit/Hyperactivity Disorder; FSIQ = Full-scale IQ.

Supplemental Table 4. Bivariate correlations between variability in hemodynamic responses to False Alarms and IIVRT.

| | Amplitude variability in hemodynamic responses to go/no-go false alarms (FA): | | | | | | | | | | | | | | | | | | | | | | | | | | | | | | Behavioral variables | | | | | |
|---|---|---|---|---|---|---|---|---|---|---|---|---|---|---|---|---|---|---|---|---|---|---|---|---|---|---|---|---|---|---|---|---|---|---|---|---|
| ICs | IC 4 | IC 6 | IC 13 | IC 14 | IC 15 | IC 17 | IC 18 | IC 21 | IC 23 | IC 28 | IC 31 | IC 33 | IC 35 | IC 36 | IC 37 | IC 39 | IC 40 | IC 41 | IC 42 | IC 44 | IC 46 | IC 47 | IC 50 | IC 53 | IC 54 | IC 55 | IC 62 | IC 64 | IC 70 | IC 71 | IC 74 | IIVRT | Mean RT | Age | ADHD | Sex | FSIQ |
| IC 1 | -0.10 | 0.32** | 0.36** | 0.19* | 0.40** | 0.39** | 0.19 | 0.27** | 0.21* | 0.28** | 0.15 | 0.31** | 0.20* | 0.19* | 0.32** | 0.15 | 0.14 | 0.24* | 0.22* | 0.09 | 0.24* | 0.11 | 0.19* | 0.19* | 0.28** | 0.15 | 0.14 | 0.18 | 0.25** | 0.05 | 0.08 | -0.08 | -0.14 | 0.00 | 0.02 | 0.16 | -0.03 |
| IC 4 | | 0.00 | -0.13 | 0.13 | -0.02 | -0.07 | 0.08 | 0.01 | 0.01 | 0.04 | -0.08 | 0.12 | 0.00 | 0.11 | -0.08 | 0.13 | 0.06 | -0.02 | -0.04 | 0.11 | 0.07 | 0.10 | -0.01 | -0.07 | 0.10 | 0.15 | 0.09 | -0.13 | 0.15 | 0.00 | 0.07 | -0.03 | -0.03 | -0.06 | 0.10 | 0.13 | 0.20* |
| IC 6 | | | 0.22* | 0.31** | 0.41** | 0.32** | 0.50** | 0.25** | 0.22* | 0.20* | 0.11 | 0.38** | 0.15 | 0.18 | 0.39** | 0.26** | 0.49** | 0.17 | 0.27** | 0.19* | 0.31** | 0.32** | 0.17 | 0.35** | 0.13 | 0.30** | 0.30** | 0.08 | 0.15 | 0.07 | 0.17 | -0.14 | -0.15 | 0.02 | -0.12 | 0.10 | -0.06 |
| IC 13 | | | | 0.11 | 0.30** | 0.23* | 0.10 | 0.23* | 0.33** | 0.32** | 0.08 | 0.17 | 0.08 | 0.25** | 0.20* | 0.23* | 0.12 | 0.29** | 0.15 | 0.22* | 0.15 | 0.09 | 0.07 | 0.29** | 0.07 | 0.21* | 0.32** | 0.13 | 0.18 | 0.08 | 0.19* | -0.03 | 0.02 | 0.01 | 0.01 | -0.06 | 0.00 |
| IC 14 | | | | | 0.21* | 0.31** | 0.31** | 0.17 | 0.04 | 0.25** | 0.23* | 0.17 | 0.07 | 0.15 | 0.20* | 0.19* | 0.22* | 0.14 | 0.12 | 0.08 | 0.11 | 0.39** | 0.10 | 0.16 | 0.24* | 0.29** | 0.28** | 0.02 | 0.24* | 0.01 | 0.04 | -0.04 | 0.10 | 0.09 | 0.02 | 0.01 | 0.02 |
| IC 15 | | | | | | 0.28** | 0.39** | 0.36** | 0.18 | 0.16 | 0.10 | 0.27** | 0.16 | 0.17 | 0.18 | 0.31** | 0.21* | 0.16 | 0.20* | 0.30** | 0.43** | 0.33** | 0.23* | 0.27** | 0.28** | 0.16 | 0.25** | 0.10 | 0.18 | 0.02 | 0.18 | -0.01 | -0.14 | -0.14 | -0.11 | 0.14 | 0.21* |
| IC 17 | | | | | | | 0.20* | 0.16 | 0.24* | 0.20* | 0.21* | 0.37** | 0.21* | 0.10 | 0.24* | 0.28** | 0.21* | 0.30** | 0.26** | 0.21* | 0.16 | 0.17 | 0.19* | 0.26** | 0.06 | 0.16 | 0.11 | 0.24* | 0.12 | -0.03 | 0.12 | -0.03 | 0.04 | -0.01 | 0.05 | 0.02 | 0.12 |
| IC 18 | | | | | | | | 0.12 | 0.21* | 0.07 | 0.08 | 0.36** | 0.01 | 0.21* | 0.22* | 0.20* | 0.34** | 0.12 | 0.23* | 0.07 | 0.31** | 0.34** | 0-.02 | 0.28** | 0.26** | 0.35** | 0.14 | 0.21* | 0.14 | 0.07 | 0.18 | -0.17 | -0.13 | -0.13 | -0.08 | 0.04 | 0.03 |
| IC 21 | | | | | | | | | 0.17 | 0.17 | 0.12 | 0.13 | 0.25** | 0.04 | 0.14 | 0.24* | 0.23* | 0.27** | 0.19* | 0.11 | 0.31** | 0.35** | 0.22* | .32** | .28** | .20* | .44** | -.01 | .12 | .17 | .18 | 0.03 | 0.12 | -0.01 | -0.09 | 0.00 | 0.00 |
| IC 23 | | | | | | | | | | 0.10 | 0.19* | 0.29** | 0.19* | 0.42** | 0.21* | 0.13 | 0.23* | 0.23* | 0.07 | 0.17 | 0.12 | 0.19* | -0.08 | 0.28** | 0.07 | 0.26** | 0.21* | 0.19* | 0.19* | 0.13 | 0.15 | -0.18 | -0.07 | 0.01 | 0.01 | -0.05 | 0.10 |
| IC 28 | | | | | | | | | | | 0.17 | 0.16 | -0.02 | 0.07 | 0.14 | 0.29** | 0.19* | 0.51** | 0.13 | 0.28** | 0.20* | 0.27** | 0.09 | 0.22* | 0.16 | 0.20* | 0.30** | 0.15 | 0.23* | 0.17 | 0.29** | -0.07 | 0.05 | 0.08 | 0.00 | 0.06 | 0.05 |
| IC 31 | | | | | | | | | | | | 0.28** | -0.09 | 0.08 | -0.02 | 0.07 | 0.11 | 0.18 | 0.07 | 0.13 | -0.11 | 0.05 | 0.19 | 0.04 | 0.04 | 0.06 | 0.17 | 0.07 | 0.20* | -0.01 | 0.17 | -0.04 | 0.05 | 0.22* | 0.15 | 0.01 | -0.04 |
| IC 33 | | | | | | | | | | | | | -0.01 | 0.02 | 0.28** | 0.30** | 0.45** | 0.27** | 0.45** | 0.31** | 0.38** | 0.19* | 0.07 | 0.28** | 0.11 | 0.10 | 0.15 | 0.27** | 0.18 | 0.21* | 0.20* | -0.11 | -0.16 | 0.01 | 0.00 | 0.01 | 0.00 |
| IC 35 | | | | | | | | | | | | | | 0.20* | 0.23* | 0.08 | 0.04 | -0.10 | 0.02 | 0.03 | 0.11 | 0.06 | 0.17 | 0.14 | 0.02 | 0.11 | 0.16 | -0.04 | 0.01 | 0.01 | -0.04 | 0.03 | -0.04 | 0.12 | 0.11 | 0.21* | -0.03 |
| IC 36 | | | | | | | | | | | | | | | 0.29** | 0.20* | 0.10 | 0.00 | 0.00 | -0.02 | 0.08 | 0.17 | 0.07 | 0.13 | 0.01 | 0.04 | 0.19* | 0.14 | 0.14 | 0.15 | 0.18 | 0.04 | 0.06 | -0.04 | 0.14 | 0.13 | 0.10 |
| IC 37 | | | | | | | | | | | | | | | | 0.12 | 0.39** | 0.22* | 0.32** | -0.05 | 0.38** | 0.14 | 0.01 | 0.44** | 0.07 | 0.11 | 0.19* | 0.12 | 0.18 | 0.02 | 0.20* | -0.03 | 0.00 | -0.13 | -0.07 | -0.03 | -0.06 |
| IC 39 | | | | | | | | | | | | | | | | | 0.22* | 0.14 | 0.08 | 0.22* | 0.29** | 0.33** | 0.30** | 0.30** | 0.11 | 0.16 | 0.39** | 0.17 | 0.13 | 0.05 | 0.10 | -0.19 | -0.10 | -0.04 | -0.07 | 0.13 | 0.17 |
| IC 40 | | | | | | | | | | | | | | | | | | 0.20* | 0.38** | 0.17 | 0.29** | 0.40** | 0.05 | 0.48** | 0.18 | 0.25** | 0.30** | 0.17 | 0.19* | 0.18 | 0.12 | -0.02 | 0.02 | 0.10 | 0.01 | 0.13 | 0.05 |
| IC 41 | | | | | | | | | | | | | | | | | | | 0.18 | 0.08 | 0.19* | 0.10 | 0.04 | 0.23* | 0.21* | 0.29** | 0.11 | 0.25** | 0.22* | 0.21* | 0.28** | -0.01 | 0.04 | -0.09 | 0.10 | -0.15 | -0.13 |
| IC 42 | | | | | | | | | | | | | | | | | | | | 0.19* | 0.30** | 0.23* | -0.01 | 0.29** | 0.06 | 0.09 | -0.01 | 0.21* | 0.22* | 0.30** | 0.08 | 0.02 | -0.06 | 0.08 | -0.04 | -0.02 | -0.08 |
| IC 44 | | | | | | | | | | | | | | | | | | | | | 0.12 | 0.18 | 0.22* | 0.30** | 0.03 | -0.04 | 0.15 | 0.18 | 0.21* | 0.13 | 0.27** | 0.01 | -0.18 | 0.09 | 0.04 | 0.14 | -0.02 |
| IC 46 | | | | | | | | | | | | | | | | | | | | | | 0.44** | 0.17 | 0.46** | 0.32** | 0.10 | 0.21* | 0.23* | 0.16 | 0.26** | 0.34** | -0.08 | -0.17 | -0.12 | -0.12 | 0.17 | 0.06 |
| IC 47 | | | | | | | | | | | | | | | | | | | | | | | 0.06 | 0.38** | 0.23* | 0.18 | 0.31** | 0.17 | 0.28** | 0.13 | 0.23* | 0.00 | -0.07 | 0.06 | -0.05 | 0.16 | 0.00 |
| IC 50 | | | | | | | | | | | | | | | | | | | | | | | | 0.13 | 0.10 | -0.03 | 0.19* | 0.10 | 0.20* | 0.05 | 0.05 | -0.04 | -0.07 | 0.13 | 0.06 | 0.22* | -0.09 |
| IC 53 | | | | | | | | | | | | | | | | | | | | | | | | | 0.06 | 0.21* | 0.36** | 0.30** | 0.25** | 0.17 | 0.31** | -0.06 | -0.08 | 0.05 | 0.01 | 0.22* | 0.02 |
| IC 54 | | | | | | | | | | | | | | | | | | | | | | | | | | 0.26** | 0.22* | 0.08 | 0.03 | 0.09 | 0.19 | 0.05 | -0.11 | -0.11 | -0.05 | 0.14 | -0.04 |
| IC 55 | | | | | | | | | | | | | | | | | | | | | | | | | | | 0.34** | 0.25** | 0.29** | 0.09 | 0.07 | -0.08 | 0.09 | -0 .01 | -0.05 | 0.04 | 0.06 |
| IC 62 | | | | | | | | | | | | | | | | | | | | | | | | | | | | 0.16 | .026** | 0.13 | 0.17 | -0.05 | 0.07 | 0.12 | 0.02 | 0.07 | 0.12 |
| IC 64 | | | | | | | | | | | | | | | | | | | | | | | | | | | | | 0.27** | 0.20* | 0.29** | -0.10 | -0.34** | 0.02 | 0.00 | 0.11 | -0.16 |
| IC 70 | | | | | | | | | | | | | | | | | | | | | | | | | | | | | | 0.31** | 0.13 | -0.15 | -0.08 | 0.20* | 0.01 | 0.11 | -0.07 |
| IC 71 | | | | | | | | | | | | | | | | | | | | | | | | | | | | | | | 0.23* | -0.10 | 0.00 | 0.15 | 0.00 | -0.03 | 0.00 |
| IC 74 | | | | | | | | | | | | | | | | | | | | | | | | | | | | | | | | -0.18 | -0.15 | 0.00 | -0.11 | 0.08 | 0.03 |

Note. ***p* <.01; **p* <.05. IC = Indenpendent Component; IIVRT = Intraindividual Variability in Reaction Times; RT = Reaction Times; ADHD; Attention Deficit/Hyperactivity Disorder; FSIQ = Full-scale IQ.

Supplemental Table 5. Bivariate correlations between variability in hemodynamic responses to Hits and IIVRT.

| ICs | IC 4 | IC 6 | IC 13 | IC 14 | IC 15 | IC 17 | IC 18 | IC 21 | IC 23 | IC 28 | IC 31 | IC 33 | IC 35 | IC 36 | IC 37 | IC 39 | IC 40 | IC 41 | IC 42 | IC 44 | IC 46 | IC 47 | IC 50 | IC 53 | IC 54 | IC 55 | IC 62 | IC 64 | IC 70 | IC 71 | IC 74 | IIVRT | Mean RT | Age | ADHD | Sex | FSIQ |
|---|---|---|---|---|---|---|---|---|---|---|---|---|---|---|---|---|---|---|---|---|---|---|---|---|---|---|---|---|---|---|---|---|---|---|---|---|---|
| IC 1 | -0.01 | 0.28** | 0.21* | 0.19 | 0.45** | 0.27** | 0.06 | 0.19* | 0.21* | 0.20* | 0.18 | 0.25** | 0.06 | 0.27** | 0.28** | 0.20* | 0.36** | 0.23* | 0.24* | 0.07 | 0.17 | 0.16 | 0.09 | 0.19 | 0.33** | 0.28** | 0.21* | 0.23* | 0.16 | 0.09 | 0.09 | -0.04 | -0.07 | 0.06 | 0.12 | 0.09 | -0.01 |
| IC 4 | | 0.16 | 0.03 | 0.16 | -0.01 | 0.07 | 0.21* | 0.11 | 0.07 | -0.03 | -0.09 | 0.15 | 0.01 | 0.08 | 0.23* | 0.10 | 0.09 | 0.02 | 0.05 | 0.19* | 0.14 | 0.31** | -0.09 | 0.10 | 0.03 | 0.08 | 0.07 | -0.06 | 0.25** | 0.11 | 0.11 | -0.06 | -0.03 | -0.08 | 0.10 | 0.08 | 0.02 |
| IC 6 | | | 0.30** | 0.20* | 0.16 | 0.30** | 0.45** | 0.18 | 0.19* | 0.13 | -0.04 | 0.32** | 0.11 | 0.21* | 0.26** | 0.06 | 0.40** | 0.10 | 0.30** | 0.12 | 0.19* | 0.31** | 0-.01 | 0.26** | 0.12 | 0.14 | 0.16 | 0.05 | 0.17 | -0.03 | 0.16 | -0.13 | -0.13 | 0.04 | -0.12 | 0.08 | -0.06 |
| IC 13 | | | | 0.25** | 0.31** | 0.17 | 0.22* | 0.14 | 0.54** | 0.30** | 0.10 | 0.07 | 0.02 | 0.28** | 0.10 | 0.11 | 0.09 | 0.18 | 0.14 | 0.15 | 0.09 | 0.26** | -0.01 | 0.19* | 0.06 | 0.22* | 0.28** | 0.10 | 0.11 | 0.17 | 0.26** | -0.04 | -0.05 | -0.01 | -0.05 | -0.02 | 0.10 |
| IC 14 | | | | | 0.16 | 0.20* | 0.25** | 0.17 | 0.12 | 0.32** | 0.13 | 0.02 | -0.03 | 0.36** | 0.31** | 0.14 | 0.33** | 0.17 | 0.13 | 0.02 | 0.25** | 0.26** | -0.03 | 0.28** | 0.19* | 0.22* | 0.26** | 0.14 | 0.09 | 0.19* | 0.33** | -0.12 | 0.03 | 0.08 | -0.10 | -0.05 | 0.07 |
| IC 15 | | | | | | 0.23* | 0.25** | 0.22* | 0.14 | 0.14 | 0.08 | 0.18 | 0.06 | 0.20* | 0.13 | 0.23* | 0.34** | 0.21* | 0.26** | 0.17 | 0.27** | 0.14 | 0.07 | 0.24* | 0.25** | 0.22* | 0.37** | 0.11 | 0.22* | 0.16 | 0.25** | 0.09 | 0.04 | -0.08 | 0.06 | 0.02 | 0.12 |
| IC 17 | | | | | | | 0.09 | 0.17 | 0.29** | 0.06 | 0.10 | 0.24** | 0.12 | 0.24* | 0.26** | 0.21* | 0.36** | 0.17 | 0.40** | 0.17 | 0.12 | 0.15 | 0.20* | 0.24* | 0.03 | 0.13 | 0.08 | 0.22* | 0.08 | -0.09 | 0.09 | -0.07 | -0.02 | -0.01 | 0.04 | 0.04 | 0.15 |
| IC 18 | | | | | | | | 0.06 | 0.26** | 0.05 | 0.07 | 0.31** | 0.02 | 0.17 | 0.18 | 0.20* | 0.38** | 0.12 | 0.29** | 0.09 | 0.22* | 0.27** | 0.02 | 0.27** | 0.28** | 0.24* | 0.23* | 0.16 | 0.27** | 0.09 | 0.24* | -0.19* | -0.22* | -0.06 | -0.03 | 0.10 | 0.10 |
| IC 21 | | | | | | | | | 0.12 | 0.01 | 0.10 | 0.15 | 0.10 | 0.07 | 0.21* | 0.14 | 0.18 | 0.23* | 0.15 | 0.04 | 0.27** | 0.22* | 0.08 | 0.19* | 0.21* | 0.13 | 0.21* | 0.05 | 0.14 | 0.13 | 0.19* | -0.02 | 0.01 | -0.06 | -0.04 | -0.01 | 0.07 |
| IC 23 | | | | | | | | | | 0.11 | 0.19* | 0.13 | -0.02 | 0.40** | 0.22* | 0.04 | 0.16 | 0.16 | 0.22* | 0.06 | 0.15 | 0.20* | -0.05 | 0.25** | 0.12 | 0.21* | 0.12 | 0.23* | 0.15 | 0.06 | 0.22* | -0.08 | -0.06 | -0.05 | 0.00 | -0.11 | 0.11 |
| IC 28 | | | | | | | | | | | 0.11 | 0.06 | -0.10 | 0.15 | 0.08 | 0.13 | 0.16 | 0.40** | 0.14 | 0.15 | 0.07 | 0.10 | -0.01 | 0.00 | 0.20* | 0.27** | 0.22* | 0.17 | 0.09 | 0.12 | 0.17 | -0.02 | 0.17 | 0.05 | -0.03 | 0.01 | 0.07 |
| IC 31 | | | | | | | | | | | | 0.07 | -0.10 | 0.08 | -0.11 | 0.13 | 0.11 | 0.21* | -0.01 | 0.00 | -0.10 | 0.06 | -0.09 | -0.09 | 0.09 | 0.21* | 0.17 | 0.07 | 0.08 | -0.06 | 0.15 | -0.08 | 0.04 | 0.21* | 0.14 | -0.08 | 0.00 |
| IC 33 | | | | | | | | | | | | | 0.03 | 0.08 | 0.21* | 0.27** | 0.39** | 0.25** | 0.46** | 0.16 | 0.15 | 0.16 | -0.04 | 0.26** | 0.07 | 0.11 | 0.16 | 0.24* | 0.15 | 0.05 | 0.03 | -0.12 | -0.16 | 0.03 | -0.03 | 0.01 | -0.01 |
| IC 35 | | | | | | | | | | | | | | 0.02 | 0.08 | 0.04 | -0.05 | -0.14 | 0.02 | 0.01 | 0.02 | -0.07 | 0.07 | 0.04 | -0.03 | 0.03 | 0.01 | -0.09 | 0.14 | 0.07 | -0.13 | -0.03 | -0.08 | 0.10 | 0.13 | 0.15 | -0.08 |
| IC 36 | | | | | | | | | | | | | | | 0.45** | 0.21* | 0.24* | 0.14 | 0.19 | 0.02 | 0.16 | 0.18 | -0.04 | 0.15 | 0.05 | 0.04 | 0.14 | 0.21* | 0.10 | 0.15 | 0.15 | 0.05 | 0.12 | -0.05 | 0.05 | 0.01 | 0.08 |
| IC 37 | | | | | | | | | | | | | | | | 0.07 | 0.34** | 0.06 | 0.37** | -0.07 | 0.33** | 0.23* | -0.05 | 0.26** | 0.13 | 0.03 | 0.26** | 0.18 | 0.19* | 0.20* | 0.08 | -0.09 | -0.04 | -0.22* | -0.14 | -0.04 | 0.01 |
| IC 39 | | | | | | | | | | | | | | | | | 0.21* | 0.33** | 0.14 | 0.18 | 0.22* | 0.12 | -0.02 | 0.15 | 0.05 | 0.13 | 0.28** | 0.24* | 0.10 | 0.09 | 0.18 | -0.16 | -0.06 | -0.03 | -0.05 | 0.16 | 0.16 |
| IC 40 | | | | | | | | | | | | | | | | | | 0.39** | 0.55** | 0.05 | 0.44** | 0.35** | -0.03 | 0.46** | 0.27** | 0.27** | 0.22* | 0.29** | 0.29** | 0.21* | 0.20* | -0.10 | -0.05 | 0.01 | -0.06 | 0.11 | 0.12 |
| IC 41 | | | | | | | | | | | | | | | | | | | 0.24* | 0.04 | 0.22* | 0.07 | 0.08 | 0.14 | 0.21* | 0.40** | 0.13 | 0.38** | 0.18 | 0.15 | 0.23* | -0.04 | 0.01 | -0.05 | 0.04 | -0.11 | -0.04 |
| IC 42 | | | | | | | | | | | | | | | | | | | | 0.14 | 0.40** | 0.22* | -0.05 | 0.29** | 0.12 | 0.07 | 0.06 | 0.32** | 0.25** | 0.26** | 0.07 | -0.08 | -0.09 | -0.01 | -0.06 | -0.04 | -0.03 |
| IC 44 | | | | | | | | | | | | | | | | | | | | | -0.05 | 0.05 | -0.02 | 0.13 | 0.08 | 0.05 | 0.12 | 0.08 | 0.08 | 0.26** | 0.13 | 0.06 | -0.06 | 0.06 | 0.09 | 0.19 | 0.00 |
| IC 46 | | | | | | | | | | | | | | | | | | | | | | 0.44** | -0.03 | 0.37** | 0.06 | 0.08 | 0.13 | 0.29** | 0.29** | 0.36** | 0.32** | -0.15 | -0.20* | -0.10 | -0.05 | 0.19 | 0.13 |
| IC 47 | | | | | | | | | | | | | | | | | | | | | | | 0.13 | 0.14 | 0.24* | 0.05 | 0.20* | 0.08 | 0.34** | 0.18 | 0.13 | 0.03 | -0.03 | -0.03 | 0.03 | 0.09 | -0.03 |
| IC 50 | | | | | | | | | | | | | | | | | | | | | | | | -0.05 | 0.09 | 0.03 | 0.10 | 0.02 | 0.07 | -0.04 | -0.01 | 0-.01 | -0.05 | -0.08 | 0.13 | -0.12 | -0.11 |
| IC 53 | | | | | | | | | | | | | | | | | | | | | | | | | 0.04 | 0.18 | 0.26** | 0.30** | 0.19* | 0.23* | 0.32** | -0.17 | -0.13 | 0.07 | -0.04 | 0.16 | 0.04 |
| IC 54 | | | | | | | | | | | | | | | | | | | | | | | | | | .21* | 0.18 | 0.09 | 0.07 | 0.09 | 0.18 | -0.03 | -0.04 | -0.11 | -0.09 | 0.04 | 0.00 |
| IC 55 | | | | | | | | | | | | | | | | | | | | | | | | | | | 0.23* | 0.34** | 0.36** | 0.10 | 0.12 | -0.11 | 0.04 | 0.06 | 0.05 | 0.01 | 0.01 |
| IC 62 | | | | | | | | | | | | | | | | | | | | | | | | | | | | 0.17 | 0.17 | 0.06 | 0.17 | -0.08 | 0.05 | 0.13 | 0.01 | -0.08 | 0.02 |
| IC 64 | | | | | | | | | | | | | | | | | | | | | | | | | | | | | 0.24* | 0.14 | 0.25** | -0.10 | -0.20* | -0.05 | 0.02 | 0.10 | 0.03 |
| IC 70 | | | | | | | | | | | | | | | | | | | | | | | | | | | | | | 0.22* | 0.06 | -0.10 | -0.10 | 0.11 | 0.13 | 0.12 | 0.04 |
| IC 71 | | | | | | | | | | | | | | | | | | | | | | | | | | | | | | | 0.17 | -0.12 | 0.00 | 0.04 | -0.03 | 0.01 | 0.05 |
| IC 74 | | | | | | | | | | | | | | | | | | | | | | | | | | | | | | | | -0.19* | -0.12 | -0.06 | -0.13 | 0.05 | 0.06 |

Note. ***p* <.01; **p* <.05. IC = Indenpendent Component; IIVRT = Intraindividual Variability in Reaction Times; RT = Reaction Times; ADHD; Attention Deficit/Hyperactivity Disorder; FSIQ = Full-scale IQ.
